# Collaborative Clinical Reasoning: a scoping review

**DOI:** 10.1101/2023.02.09.23285741

**Authors:** Ching-Yi Lee, Hung-Yi Lai, Ching-Hsin Lee, Mi-Mi Chen, Sze-Yuen Yau

## Abstract

**Introduction:** Team approaches in healthcare highlight the importance and benefits of collaboration on maximising clinical outcomes and patient safety. Based on extant literature, the authors aimed to explore collaborative clinical reasoning (CCR) among healthcare professionals.

**Methods:** A scoping review was undertaken to examine CCR related studies in healthcare. Medline, PsychInfo, SciVerse Scopus, and Web of Science were searched. Inclusion criteria included full-text articles published between 2011 to 2020. Search terms included cooperative, collaborative, shared, team, collective, reasoning, problem solving, decision making, combined with clinical or medicine or medical, but excluded shared decision making.

**Results:** A total of 24 articles were identified in the review. Analysis of the articles resulted in four major content themes: (1) Decision-making process (n=14); (2) Quality assessment by MDTs-MODe (Multidisciplinary Team-Metric for the Observation of Decision Making; n=5) (3) CCR theory and definitions(n=3); and (4) Problem-solving process (n=2). Most articles focused on communication associated with collaborative decision-making processes. The discussion of team impacts among all studies was merely the notion of clinical reasoning as an essential component of the collaborative or interprofessional practice. None provided direct evidence on the process of CCR or its impact on clinical outcomes. Only two articles provided specific definitions on CCR.

**Discussion:** We illuminate the necessity of further research in CCR, specifically with a focus on cognitive components of CCR. A better understanding of current CCR research in healthcare may inform future discussions around establishing strategies to enhance CCR development, and hence provide positive influence on medical education and patient safety.

## Introduction

### Clinical Reasoning Errors

To err is human [1], a normal human behavior places every health professional at risk of unexpected medical errors generated from misdiagnosis or a minor mistake. Many researchers have attempted to quantify the relationship between wrong diagnoses and increased morbidity and mortality [2, 3]. Diagnostic errors have been recognized as a primary cause of death in the United States, with an estimated error rate of 10% to 15% according to the autopsy data [4, 5]. Although the occurrence of errors only takes up a minority of all cases, the majority of errors that do occur result, at least in part, from the individual doctor’s cognitive processes [6]. Faulty clinical reasoning is considered one attribution to diagnostic errors, and several studies have indicated that resolving the issue of error prevention requires an improvement in clinical reasoning skills [2, 7, 8].

### Clinical Reasoning in Medical Education

Clinical reasoning is “the thought process that guides practice” and it is a central component of physician competence [8]. Other related terms for clinical reasoning include problem-solving, decision-making, critical thinking, and judgment [9]. The term clinical reasoning has been described as the process by which clinicians collect cues, process the information, come to an understanding of a patient problem or situation, plan and implement interventions, evaluate outcomes, and reflect on and learn from the process [10]. One of the influential models used to describe the details of clinical reasoning process, namely “dual-process’ theories of cognition, argued that errors are usually associated with “system 1” thinking (instant, automatic and intuitive mode of thinking), originated in cognitive heuristics [11]. Although system 1 thinking shows the advantage of developing instant judgments by pattern recognition, it is susceptible to the associated biases and the impact of the emotional content of the person who makes such judgment [12]. And therefore, the process of current information primarily relies on the previous scenarios or experience stored in memory which may somehow leads to incomplete or wrong diagnosis.

Another extreme of dual-processing theorists argued errors are related to forced “system 2” thinking (slow, effortful and analytic mode of thinking) [6, 13]. When appropriate data are available system 2 yields the most normatively national reasoning, but it is easily disrupted by high cognitive load [12]. Conscious reasoning (i.e. system 2 thinking) places a heavy workload on cognitive resources, especially on working memory. Working memory provides a workplace for maintaining and manipulating temporary information. It is a limited resource and is competed for by both processing and storage tasks [14]. If a task’s demand exceeds one’s capacity, it may not be possible to maintain the information stored in working memory or processing disrupted, which results in errors.

### Interprofessional Collaborative Clinical Reasoning

Previous collaborative healthcare literatures largely focused on the teamwork competencies and interprofessional collaboration [15–17]. The conventional discussion of team impacts on healthcare professional competences primarily focused on individualist discourse. They emphasized on the outcomes, with the individual gain that practitioners acquire, perform, and maintain over their practice life. The notion of “collective competencies” shed light on the underlying mechanism of teamwork [18]. It addresses how individually “incompetent” healthcare professionals shared and distributed to form a “competent” team. This collectivist discourse focuses on the similarities and differences that each practitioner perceived in the situation, and how they trigger and share the mental models among the various team members. The term “collaborative reasoning” proposed by Mason will be employed to describe the process of reaching a shared mental model [19]. It was proposed that participants in a team could sort out the solution more effectively and efficiently by anticipating with other members’ responses. Furthermore, the complexity of the situation was positively correlated with the influential effect of shared mental model. One of the insights was that the degree to which team members shared is positively correlated with the team performance [20].

### Significance of Current Study

Both systems 1 and 2 thinking in dual-processing model of clinical reasoning are constantly adopted and interchangeable during decision-making among healthcare professionals. However, it is generally recognised that system 1 thinking favoring pattern-recognition costs less effort than system 2 thinking which demands more mental searches. In a busy clinical setting, it is impractical for an individual healthcare professional to stay in system 2 thinking continuously, however, this type of thinking is often crucial and less prone to error [12, 13, 21]. Collaborative clinical reasoning, a process similar to the idea of shared mental model may allow cognitive load sharing in a complex clinical situation where multiple healthcare professionals are involved. It may help identify, reduce subjective biases and leads to efficient decision-making during diagnostic processes through team effort and communication [17–20]. Presently, only few studies have explored collaborative clinical reasoning among healthcare professionals, specifically the process and factors associated with this practice is seldom discussed. Since a scoping review enables one to identify and map the types of available evidence in a given field [22], we believe there to be more related evidence should we examine the field under close scrutiny. Given that recent literature review in clinical reasoning reported only a limited number of studies relating to collaborative performance [23], and that only nine articles with at least one physician involved were found in the review, a scoping review specifically focusing on collaborative clinical reasoning involving any particular healthcare professional team was therefore employed in this study.

## Method

In accordance with the Arksey and O’Malley framework [24], and the recent recommendations by Levac et al., [25] the framework of this methodology involves the following steps: (1) scoping review questions, (2) search strategy, (3) study screening and selection, (4) data extraction, and (5) analysis and presentation of results.

### (1) Review questions

This review is guided mainly by the question, “What is the current status of collaborative clinical reasoning (CCR) research in general?”. After discussion by the research team, more specific research questions were further developed: (1) How many CCR papers were published over the decade? (2) What is the theory and methodology used in these published journals? (3) What were the main populations being studied? (4) Which topics were most frequently published? and (5) Are there any trends over time?

### (2) Relevant studies and search strategy

The initial search involved four electronic databases: Medline, PsychInfo, SciVerse Scopus (multidisciplinary, 1823-present), and Web of Science (multidisciplinary, 1900-present). We limited the search to the past ten years (2011-2020) to identify the latest trends of current researches. The language of articles is limited to English. Using Kiesewetter’s search strategy [23], the search terms include cooperative, collaborative, shared, team, collective, reasoning, problem solving, decision making, combined with clinical or medicine or medical, but exclude shared decision making. The primary interest of subjects were associated only with healthcare professionals who were involved actively in clinical activities. The studies involving patients or trainees such as students and interns were excluded.

### (3) Study selection and screening

All papers were collected and managed using EndNote® software to eliminate duplicates. Initially, only the title and abstract of citations would be screened independently by two reviewers to preclude waste of resources in procuring articles that fail to meet the minimum inclusion criteria. The exclusion criteria were applied to non-peer-reviewed paper, conference, letters or editorial articles, papers lack of original data, and those without full-text available. Papers involved discussion mainly about individual clinical reasoning itself but without any types of team effort or collaborative interaction were excluded.

### (4) Data charting

After the title and abstract screening process, all papers considered relevant were imported to ATLAS.ti™ from EndNote®. A charting content was developed using ATLAS.ti™ to confirm relevance and to extract study characteristics such as publication year, publication type, study sector, terminology, methodology, number and types of research participants. This charting process was reviewed by the research team and pretested by all reviewers before implementation. The characteristics of each full-text article were extracted by two independent reviewers. Studies failing to meet the eligibility criteria were excluded further in this phase. Reviewers met throughout the process to resolve conflicts and ensured consistency with the research questions and purpose.

### (5) Data summary and synthesis

An analytical framework of quantitative and thematic approach was used to collate various themes that emerged from the existing data. An overview of basic descriptive frequency counts was presented instead of synthesizing results of the included studies. Therefore, article ‘demographic’ coding such as year or journal, and thematic coding of content were used to report the data. Frequencies of counts were summarized and presented in graphical or tabulated form. Microsoft Excel 2010 (Microsoft, Redmond, WA, USA) was used to facilitate descriptive analyses and graphical summaries. Each article was coded by a maximum of two themes during the coding analysis by ATLAS.ti™. The charting results were then summarized using Microsoft Excel 2010 (Microsoft, Redmond, WA, USA) and percentages were utilized to describe the nominal data.

### (6) Team consultation

The research members met on a weekly basis to track the progress of the scoping review, and monthly meetings were held with the international consultant for further consolidation of results.

## Results

The searches initially retrieved a total of 281 citations from the four of the databases specified earlier. Following the database searches, snowballing through hand searching reference lists results in a total of 1 additional article. After the duplication check, 134 citations remained for the first screening. Titles and abstracts were screened, yielding 89 records of articles considered eligible for full review. A further 65 records were excluded because they were articles identified as editorial, thesis or book (10); and articles without original data (1) or content related to collaborative clinical reasoning (40) or healthcare professionals involved (14). Following data characterization, 24 articles were included for full coding (see Figure S1 in the Electronic Supplementary Material, ESM).

### Year, Journal, and Methodology (RQs 1–2)

The number of annually published articles on CCR alternates at values of 1 or 2 between 2011 and 2016 (Figure 1). The annual sum increased to 3 in 2017 but declined to 1 again in 2018. The highest and second highest number of CCR studies were found in 2019 (n=6) and 2020 (n=4), respectively. The journals with which these 24 articles were published were listed alphabetically in Table S1, found in ESM. There were only 2 articles published in the same journal, Annals of Surgical Oncology. Each journal as suggested by its name was categorized into six genres. The majority of the articles fell into categories of oncology (n=8) and medicine in general (n=7) while the rest of the articles made up the categories of nursing (n=2), medical education (n=3), ergonomics or medical informatics (n=2), and philosophy or psychology (n=2). Both quantitative (n=11) and qualitative (n=10) methodology were the most prevalent approaches while mixed methods (n= 3) was the least common approach.

**Figure 1.**
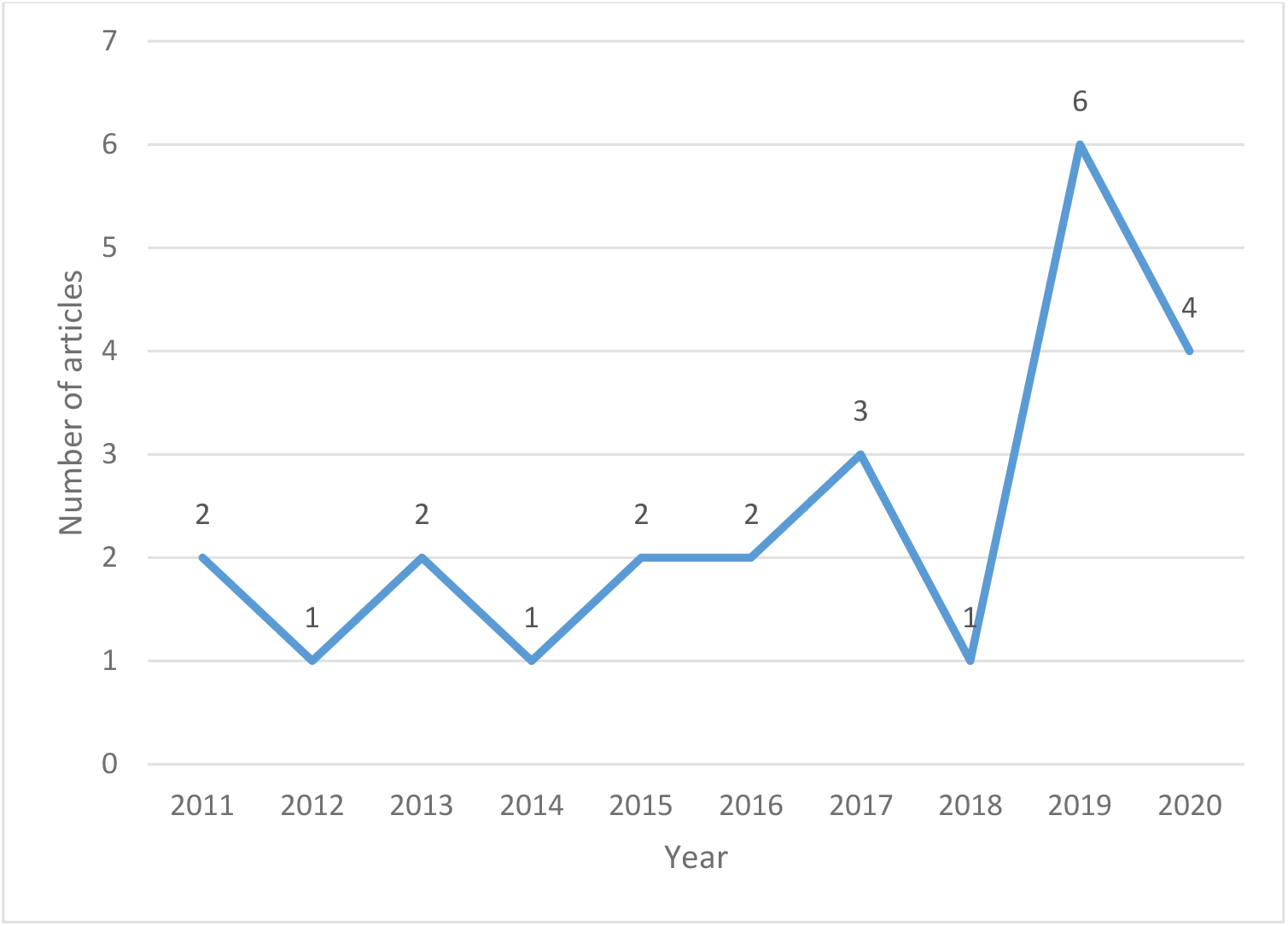
Annual number of articles on collaborative clinical reasoning between 2011 and 2020.

### Themes, Population, and Trends (RQs 3–5)

In table 1, the matching of the articles into four major content themes were generated in a decreasing order based on the frequency in our data, namely: (1) Decision-making process (n=14) [26–39]; (2) Quality assessment by MDTs-MODe (Multidisciplinary Team-Metric for the Observation of Decision Making; n=5) [40–44](3) CCR theory and definitions (n=3) [23, 45, 46]; and (4) Problem-solving process (n=2) [47, 48]. The double coding frequency was also used to support the key issues identified within our content themes.

**Table 1.** Content themes for articles on collaborative clinical reasoning between 2011-2020.

| Major Content Themes | Single Coding Frequency |
| --- | --- |
| **Decision-making process**<br>Any article directly addresses the topic of “decision-making” process in the title or keyword, or as the subject of interest throughout the context.<br>[26-39] | 14 |
| **Quality assessment by MDTs-MODE**<br>Articles involve the assessment of multi-disciplinary team meetings (MDT or MDM) using the standard MDT-MODE (Multidisciplinary Team-Metric for the Observation of Decision Making).<br>[40-44] | 5 |
| **Collaborative clinical reasoning theory and definitions**<br>Articles specifically explain the theory or definitions about collaborative clinical reasoning.<br>[23, 45, 46] | 3 |
| **Problem-solving process**<br>Articles directly address the topic of “problem solving” process in the title or keyword, or as the subject of interest throughout the context.<br>[47, 48] | 2 |
| Minor Content Themes | Double Coding Frequency |
| Articles involve multi-disciplinary team meetings (MDT or MDM) (eg. 13 cancer MDTs, 1 thoracic) | 14 |
| Communication and other factors (eg. culture) in decision making | 8 |
| Collective intelligence (eg. compositional team cognition) | 4 |
| Trigger for decision-making (eg. Nurses initiate decision-making) | 2 |
| Team conversational data (their relation to decision-making or problem solving) | 2 |
| Simulation in ward | 1 |

#### I. MDT Participants and Data Collection

Overall, there are 14 articles involving studies conducted with MDT members [26–28, 30, 34, 36, 39–44, 47, 48]. Only 1 article among these MDT-related articles collects both non-cancer and occasionally cancer related MDT data in a thoracic ward [36]. The remaining 13 articles all address issues about cancer MDT, 5 of which focus on MDT quality assessment utilising the tool, MDTs-MODe. The most discussed MDT case was colorectal or gastrointestinal cancer. In terms of the MDT composition, nurses or nurse specialists were the most frequently identified team member. The second and third highest propotion of team members, namely surgeons, radiologists, histopathologists and oncologists entails how they are often coupled with nurses or nurse specialists, and altogether they often represent the common composition of team members found in a cancer MDT.

#### II. Non-MDT-specific articles

These studies do not specifically include the term MDT, however there are few of them do fall into the category of team concept. These studies are also summarized by minor themes (Table 1). Two reviews describe the theory about CCR [23, 46] while one review characterizes collective intelligence in medical decision making [35]. Two comparative studies show evidence on better performance in teams than individuals when solving a cognitive drug problem [33] or interpreting mammograph screening [32]. One study qualitatively compares the different decision-making process on antibody prescriptions between emergency and surgical teams, where the authors attribute such difference to team culture [37]. One simulation study conducted with residents and nurses in internal medicine wards identifies characteristics and defines 5 dimensions of CCR [45]. Two studies demonstrate the importance of communication during DM process, and specifically the role of a nursing staff on initiating a DM process in a team [29, 38]. Upon qualitative analysis of informal conversations about patient cases in a medical team, one study reveals three collaborative practices: (a) joint interpretation, (b) intersubjective generation and validation of hypotheses, and (c) postponing the diagnostic decision [31]. In general, several articles have addressed separately how communication, trust, team composition, institutional culture, or prescriptive authority may exert an influence on collaborative practice in healthcare team decision-making [28–31, 33, 34, 36–39, 45, 48].

## Discussion

This scoping review of CCR research identified a resultant of 24 studies published between 2011 and 2020. Decision-making process was the most dominant theme found across these studies. A majority of the articles focus on communication and factors associated with collaborative decision-making processes. The discussion of team impacts among all studies was merely the notion of clinical reasoning as an essential component of the collaborative practice or interprofessional development. Moreover, there were not many articles which provide details such as the structure, process or definition of CCR. In depth, only two studies provide a more specific and relevant definitions on CCR [23, 45]. Kiesewetter et al. summarized factors that may influence the performance of CCR: (1) The initial distribution of information, (2) practitioners’ clinical experience in a team, (3) information exchange among members, and (4) individual retrieval and representation of the information that shared by a team such as distribution of information or clinical experience [23]. In a simulation study conducted in health care setting, Blondon et al. have identified five dimensions of collaborative reasoning in internal medicine: (1) diagnostic reasoning, (2) patient management, (3) patient monitoring, (4) communication and (5) explanations to patient [45]. Based on the definition of CCR from these two studies [23, 45], one review emphasizes the importance of clinical reasoning collaboration in relation to the development of shared decision-making or inter-professional education [49]. In this review, the authors cited two literature using conventional clinical reasoning learning methods such as case study framework for training healthcare professional students but without exploring details on cognitive components of CCR [49].

The literature search of collaboration in healthcare has been identified with the following terms: interdisciplinary, multidisciplinary, interprofessional and intraprofessional. In general these terms are often described interchangeably with teamwork, team approaches, collaborative practice, coordination and cooperation [50–53]. Currently, the concept of team approaches has been advocated over the past decade by various healthcare settings such as internal medicine or trauma care [54–56]. Several studies have addressed the benefits of collaboration practices on maximising patient safety and clinical outcomes [57–61]. Only a handful of interprofessional related studies were found to include discussion on clinical reasoning [23, 45, 46, 49, 62, 63]. Muller-Juge et al. conducted semi-strucutred interviews with nurses and residents investigating their role perceptions and expectations on interprofessional collaboration in an internal medicine ward [62]. In their study, one of the thematic findings revelaed that both professions perceived that the residents play the major role of performing clinical reasoning [62]. In the same setting of internal medicine and semi-strucutred interviews, the nurses and physicians from another study by Wölfel et al., regarded CCR as core-competences and particularly important for interprofessional development [63]. However, in strict terms, these two studies showed rather little relevancy to CCR despite clinical reasoning was mentioned as an essential component of the collaborative practice.

In the descriptive article by Olson et al., team clinical reasoning observed at the existing healthcare setting often leads to a “parallel play” instead of the actual collaborative practice [46]. To overcome such issue, attention has been placed on team communication in order to enhance information exchange and optimize decision-making during collaborative practice [64, 65]. Although communication is one of the dimensions identified during the process of CCR [45], another dimension such as diagnostic reasoning is seldom found across the literature. This is also reflected in our scoping review results where a group of MDT studies demonstrate the use of quantitative MDT-MODe to assess information retrieval and communication among healthcare teams to evaluate the quality of decision making.

This review has identified current trends of research areas on CCR. With respect to the major code and the theory based on Donabedian’s model [66], the process component of CCR, decision-making process, and factors associated with decision-making such as communication are the most dominant themes among the literature found in this review. The second mostly discussed topics of interest are related to the structure and outcomes of CCR. First, the most common setting involves cancer teams. As mentioned earlier, most CCR studies involve occasions of cancer MDT meetings (MDMs) and therefore its member structure often includes healthcare professionals specific to the target cancer for team treatment. As a result, the outcomes of CCR are frequently associated with the quantitative assessment of MDT-MODe. Given that CR is a cognitive process that is challenging to measure and evaluate [8], it is anticipated that most studies on CCR take a relative approachable form of this quantitative measure.

### Implication

With extensive experience and years of accumulated knowledge, expert physicians can categorize and process relevant information of diseases quicker, since they have superior schema and pattern recognition skills [67]. In terms of dual-process model in clinical reasoning, such act referring to system 1 thinking has potential risks of making errors in diagnosis. Team communication related to CCR may help team members stay abreast of each other’s thoughts and improve their efficiency by elimination of individual biases [45]. On the other hand, conscious reasoning associated with system 2 thinking is prone to place a heavy workload on cognitive resources and hence increase the likelihood of errors [14] [68]. In a healthcare team, cognitive resources could be shared among the professionals to efficiently process clinical tasks. Each individual is responsible for manipulating and transforming information within their capacities and sharing the intermediate results with members. For example, it is anticipated that the senior physicians may make reinterpretation based on additional investigation of findings given by other team members. This process of reflection allows retrieval of new information stored in memory, leads to the development of different working hypotheses and thus a new interpretation [69].

Evidence from aviation and aerospace literature suggests that a team-based competency enhances both coordination and productivity, which can also be applied to healthcare settings [70]. The term “collaborative reasoning” describes the process of reaching a shared mental model by working with team members [71]. Participants in a healthcare team can arrive at a solution more effectively and efficiently by anticipating other members’ responses and needs. In situations involving a greater degree of complexity, the value of having a shared mental model becomes more apparent. Indeed, the extent to which team members contribute to shared effort correlated positively with team performance [72]. In congruence with the recent review, it has shown the ability of the group to outperform an individual on cognitive tasks, and where evidence also suggests that team-based care has important implications for medical diagnosis and decision-making [35]. Given that team-concept has become a popular approach to the diagnostic process in the provision of quality health care, it is important to promote fundamental research in collaborative reasoning in healthcare teams in order to fully understand the underlying mechanism and specifically cognitive components of clinical reasoning processes during collaborative practice.

## Conclusions

This study provides the literature overview on CCR research. Limited evidence has demonstrated how team reasoning performance in a clinical encounter may be influenced by factors such as communication. Further insight into the cognitive process, the diagnostic dimension of collaborative clinical reasoning is apparently essential. An awareness of the cognitive processes in CCR shall inform medical educationalists in refinement of further strategies to enhance CCR development, and hence potentially increase a healthcare professional’s ability to provide safe and effective patient care.

## Data Availability

All data produced in the present work are contained in the manuscript

## Additional information

Ching-Yi Lee and Hung-Yi Lai contributed equally to this work and are co-first authors.

## Supplementary Information

**Figure S1.**
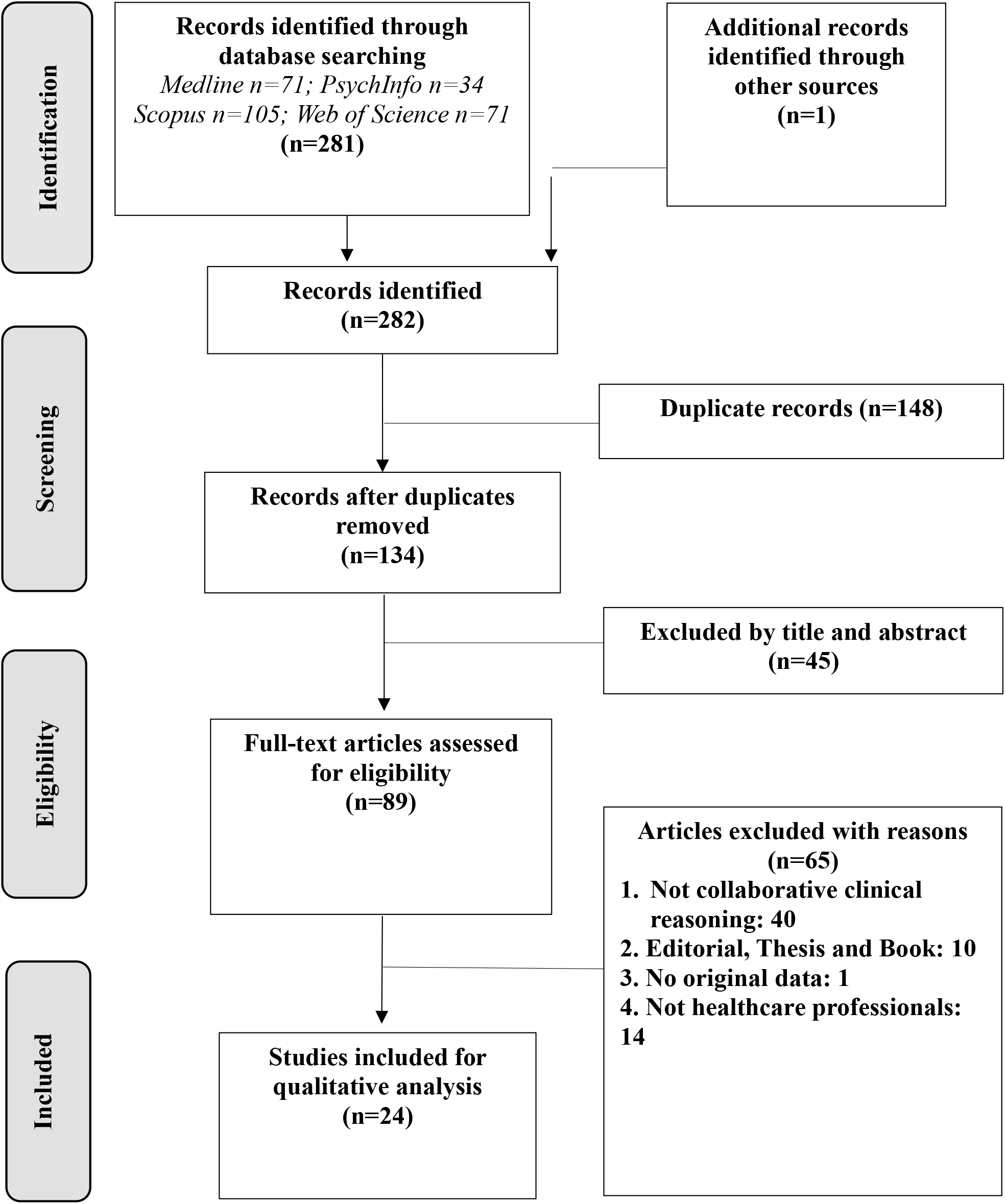
Flowchart of the study selection process.

**Table S1.** Categorisation of 24 collaborative clinical reasoning articles into journal genres.

| **Genres**<br>**Journals** | **Oncology** | **Nursing** | **Medical**<br>**Education** | **ERGONOMICS**<br>**or**<br>**Medical**<br>**Informatics** | **Psychology**<br>**&Philosophy** | **Medicine**<br>**in general** |
| --- | --- | --- | --- | --- | --- | --- |
| Annals of Surgical<br>Oncology | 2 |  |  |  |  |  |
| Behaviour &<br>Information<br>Technology |  |  |  | 1 |  |  |
| BMC Cancer | 1 |  |  |  |  |  |
| BMC Medical<br>Informatics &<br>Decision Making |  |  |  | 1 |  |  |
| Cancer Medicine | 1 |  |  |  |  |  |
| Clinical Infectious<br>Diseases |  |  |  |  |  | 1 |
| Communication and<br>Medicine |  |  |  |  |  | 1 |
| Diagnosis |  |  |  |  |  | 1 |
| European Journal of<br>Cancer Care | 1 |  |  |  |  |  |
| European Journal of<br>Oncology Nursing | 1 |  |  |  |  |  |
| International Journal<br>of Surgery |  |  |  |  |  | 1 |
| Journal of Advanced<br>Nursing |  | 1 |  |  |  |  |
| Journal of Clinical<br>Nursing |  | 1 |  |  |  |  |
| Journal of<br>Continuing<br>Education in the<br>Health Professions |  |  | 1 |  |  |  |
| Journal of Geriatric<br>Oncology | 1 |  |  |  |  |  |
| Medical Teacher |  |  | 1 |  |  |  |
| Medicine |  |  |  |  |  | 1 |
| Mind Culture and<br>Activity |  |  |  |  | 1 |  |
| Oncology Research<br>and Treatment | 1 |  |  |  |  |  |
| Plos One |  |  |  |  |  | 1 |
| Postgraduate<br>Medical Journal |  |  | 1 |  |  |  |
| Synthese |  |  |  |  | 1 |  |
| World Journal of<br>Surgery |  |  |  |  |  | 1 |
| **Total** | **8** | **2** | **3** | **2** | **2** | **7** |

